# Assessment of childhood vaccine immunization coverage, card retention, and compliance in Bayelsa State post-Gavi support: A household survey of children aged 0-59 months

**DOI:** 10.1101/2025.08.11.25333409

**Authors:** Ebiakpor Bainkpo Agbedi, Mordecai Oweibia, Christopher Peres Ekiyor

## Abstract

**Background:** Immunization is a vital public health intervention for preventing childhood diseases and reducing mortality rates. Despite global efforts, Nigeria, particularly Bayelsa State, continues to experience suboptimal immunization coverage, often exacerbated by logistical challenges, caregiver misconceptions, and cultural beliefs. The GAVI Alliance has aimed to enhance vaccine availability and community engagement in immunization programs, yet the impact on local coverage and card retention remains under-researched.

**Objective:** This study aimed to assess the current immunization coverage, card retention, and barriers to vaccination among children aged 0-59 months in Bayelsa State, following GAVI support.

**Methods:** A descriptive cross-sectional survey was conducted, utilizing a multistage sampling approach to collect data from 420 caregivers of eligible children through structured interviews and immunization card verification. Data were analysed using descriptive and inferential statistics, including Chi-square tests and logistic regression.

**Results:** The findings revealed an overall immunization coverage of 95.5%, a significant increase from the baseline of 61.12%. BCG and OPV vaccines demonstrated high coverage rates (93.6% and 94.3%, respectively), while the measles and yellow fever vaccines showed lower coverage (56.0% and 54.5%). Notably, caregiver concerns about vaccine safety and potential side effects were prevalent, influencing non-compliance, with 34.6% citing forgetfulness as a reason for missed vaccinations. Moreover, educational level and distance to health facilities were significant factors affecting immunization card retention and compliance.

**Conclusion:** The study underscores the positive impact of GAVI-supported initiatives on immunization coverage in Bayelsa State. However, persistent gaps in specific vaccine uptake and caregiver perceptions necessitate targeted educational interventions and improved access to healthcare services to sustain high immunization rates and protect children from vaccine-preventable diseases.

## 1.0 Introduction

### 1.1 Background

Immunization is globally recognized as one of the most effective interventions for preventing childhood diseases and reducing child mortality rates. According to the World Health Organization (WHO, 2019), immunization prevents approximately 2-3 million deaths each year from diseases such as measles, diphtheria, pertussis, and tetanus. Achieving high immunization coverage is essential for controlling vaccine-preventable diseases and ensuring healthy development in children. In Nigeria, despite national efforts and international support, immunization coverage remains suboptimal. The 2018 Nigeria Demographic and Health Survey (NDHS) reported that only about 31% of children aged 12-23 months were fully immunized—far below the global target of 90% coverage set by the WHO (NDHS, 2018). Factors contributing to this low coverage include inadequate health infrastructure, logistical challenges, caregiver misconceptions, cultural beliefs, and issues related to vaccine supply and cold chain management (Wiysonge CS, 2012; Williams SV, 2024). To address these challenges, the GAVI Alliance (GAVI, 2018) has partnered with Nigeria to support immunization programs through funding, vaccine procurement, capacity building, and community awareness initiatives. Since the implementation of GAVI-supported interventions, there have been notable improvements in vaccine availability, outreach activities, and immunization coverage, especially in underserved and hard-to-reach communities. However, the success of these programs depends not only on vaccine availability but also on community acceptance, caregiver participation, and effective record-keeping. Immunization cards serve as critical tools for tracking individual vaccination histories, monitoring coverage, and evaluating program impact. Loss or non-use of immunization cards can hinder accurate assessment and lead to missed doses or unnecessary re-vaccination (Ogbuagu I, 2024). Despite these efforts, recent data specific to Bayelsa State are limited, and there is a need for community-level assessments to evaluate the current immunization status, identify barriers, and inform targeted interventions. Understanding caregiver knowledge, attitudes, and practices is essential for designing culturally appropriate strategies to improve vaccine uptake. Therefore, this study aims to evaluate the current immunization coverage, card retention, and barriers to vaccination among children aged 0-59 months in Bayelsa State. The findings will provide evidence to guide policymakers, health workers, and stakeholders in optimizing immunization strategies and achieving national and global immunization goals.

Vaccine-preventable diseases (VPDs) are illnesses that can be prevented through the administration of vaccines. These diseases have been a major cause of morbidity and mortality worldwide, particularly among children. VPDs pose a significant threat to global health, especially among vulnerable populations such as children and the elderly. Diseases like measles, polio, diphtheria, pertussis, and tetanus have resulted in millions of deaths and disabilities globally. According to the World Health Organization (WHO), vaccine-preventable diseases are estimated to claim the lives of over 1.5 million children under the age of five each year. Besides mortality, VPDs can lead to long-term health consequences, such as disabilities, developmental delays, and increased susceptibility to other infections. Several factors contribute to the persistence of VPDs. Inadequate access to vaccines, particularly in low-income countries, can result in low vaccination coverage, facilitating the spread of diseases. Misinformation and misconceptions about vaccine safety and efficacy can lead to vaccine hesitancy, further reducing vaccination rates and increasing outbreak risks. The increased mobility of people and goods across borders also facilitates the transmission of VPDs. Vaccination remains the most effective method to prevent VPDs. Immunizing individuals against specific diseases reduces the risk of illness and death and can prevent outbreaks by decreasing the number of susceptible individuals in a population. High vaccination coverage can protect those who are not vaccinated, such as individuals with weakened immune systems, by providing herd immunity. To combat VPDs, it is essential to implement effective strategies that engage communities, healthcare providers, and policymakers. The formulation and implementation of culture-sensitive strategies enhance and strengthen immunization programs, which are crucial for preventing VPDs. This can be accomplished by improving vaccine accessibility, ensuring that vaccines are readily available to all individuals, particularly in low-income and hard-to-reach populations. Implementing efficient vaccine delivery systems, such as mobile vaccination sessions and community-based vaccination programs, is vital for preventing outbreaks of vaccine-preventable diseases. Providing healthcare providers with the necessary training and resources to administer vaccines effectively and safely is paramount in achieving protection against these devastating diseases. Community engagement and education are vital for promoting vaccination and increasing service uptake of immunization. Public awareness campaigns can educate communities on the importance of vaccination and the risks associated with VPDs. Engaging with stakeholders, including paramount chiefs, local leaders, influencers, and the broader community, is essential to promoting vaccination. Developing targeted strategies to address vaccine hesitancy, particularly by tackling misconceptions and misinformation, is key. Robust surveillance systems and outbreak response mechanisms must be established to detect and respond to VPD outbreaks promptly. Developing plans for responding to VPD outbreaks, including rapid vaccination campaigns and contact tracing, alongside collaboration between governments, international organizations, and civil society organizations, is essential for preventing VPDs. Immunization is a cornerstone of public health, protecting millions worldwide from vaccine-preventable diseases. However, immunization strategies face challenges that hinder their effectiveness. Funding constraints pose a significant challenge; reduced donor funding has led to moderate to severe disruptions in vaccination campaigns, routine immunization, and access to supplies in nearly half of the countries surveyed by WHO. This funding crisis severely limits the ability to vaccinate vulnerable children in fragile and conflict-affected countries. Misinformation about vaccines can lead to vaccine hesitancy, which reduces vaccination rates and increases the risk of outbreaks. The spread of misinformation through social media has complicated efforts to address vaccine hesitancy and promote immunization. Population growth, humanitarian crises, and conflicts also present significant challenges to immunization strategies. These situations can lead to population displacement, destruction of healthcare infrastructure, and reduced access to vaccines and immunization services, consequently putting millions of children and adults at risk of vaccine-preventable diseases. Recent outbreaks of VPDs, such as measles, meningitis, and yellow fever, are on the rise globally, with diseases like diphtheria at risk of re-emerging. These outbreaks emphasize the need for sustained investments in vaccines and immunization programs, as well as efforts to improve global vaccination coverage. Immunization services, disease surveillance, and outbreak responses in nearly 50 countries are already experiencing disruptions similar to those seen during the COVID-19 pandemic. Such disruptions can lead to lower vaccination rates, increased outbreak risks, and a decreased ability to respond to emerging health threats. Low routine vaccination coverage and gaps in preventive campaigns elevate the risk of outbreaks, particularly in regions with no reported cases in the past, underscoring the need for targeted interventions to enhance vaccination coverage. Despite its benefits, vaccine hesitancy remains a growing concern globally. The WHO has identified vaccine hesitancy as one of the top ten threats to global health, emphasizing its multifactorial nature, which includes complacency, confidence, and convenience. Understanding the underlying factors influencing vaccine hesitancy is crucial for designing effective interventions. Despite significant global efforts and substantial reductions in polio cases since the launch of the Global Polio Eradication Initiative in 1988, Nigeria remains one of the few countries where the disease is still endemic, particularly due to issues surrounding vaccination compliance. A study conducted by Jibrin AM (2025) investigates the multifaceted drivers of non-compliance with polio vaccination programs across four states in Nigeria: Kano, Taraba, Edo, and Abia. This research highlights the complex interplay of socio-cultural, economic, institutional, and informational factors contributing to vaccine hesitancy and refusal in Nigeria. A significant finding of the study is the widespread lack of knowledge regarding polio and its vaccination program among caregivers, with over 68% of respondents demonstrating inadequate awareness. The study reveals that 35.4% of respondents expressed concerns about the potential side effects of the polio vaccine, including fears of infertility. Furthermore, a notable percentage of individuals indicated that religious teachings and community leaders significantly impact their vaccination decisions, with 47.6% acknowledging religious influences that either support or discourage vaccine acceptance. This finding aligns with existing literature emphasizing the role of socio-religious narratives in vaccine refusal, particularly in northern Nigeria. A systematic review by MacDonal et al. (2015) synthesized evidence on vaccine hesitancy, identifying key drivers such as misinformation, socio-cultural beliefs, and trust in healthcare systems. This aligns with findings from Agbede et al. (2024), which emphasized the influence of cultural beliefs and religious practices on vaccination uptake in Nigeria, suggesting that cultural contexts significantly shape parental attitudes toward immunization. Research has shown that socio-demographic variables play a critical role in shaping vaccine-related behaviours. For instance, a study conducted in sub-Saharan Africa revealed that lower levels of parental education were associated with higher rates of vaccine refusal and non-compliance (Wiysonge CS, 2012). Higher levels of parental education correlate with increased adherence to vaccination schedules, as educated parents are often more informed about the benefits of immunization and less susceptible to misinformation (Dube’ et al., 2016). Conversely, low educational levels among parents, as found in various studies, are linked to heightened vaccine hesitancy and refusal, emphasizing the critical need for targeted educational interventions (Jibrin AM, 2025). This is echoed by others, who found that parents with limited education are less likely to understand the importance of vaccines, leading to higher rates of zero-dose children in their communities (Patrick B, 2025). The present study similarly identifies educational status as a pivotal determinant influencing immunization rates in northern and southern Nigeria. Additionally, the role of household dynamics, particularly the gender of the household head, has been highlighted as a significant factor influencing health-related decision-making. Studies have indicated that in patriarchal societies, where males predominantly occupy household leadership roles, there may be disparities in healthcare access and utilization, ultimately affecting children’s immunization status (Hossain KAHM, 2021). The findings from the current study reinforce this notion, revealing that households led by females exhibited a markedly lower immunization rate compared to those led by males. Further studies have highlighted that female-headed households often face additional barriers, such as reduced mobility and authority in decision-making, which can hinder access to healthcare services (Tracey G, 2024). Similarly, this research illustrates that households with male heads are associated with higher immunization rates, emphasizing the need for gender-sensitive health interventions. Furthermore, the role of communication and information dissemination is critical in addressing vaccine hesitancy. There remains a notable reliance on informal sources of information, such as community rumours, which can perpetuate misinformation. This reliance on unofficial channels can undermine the effectiveness of health messages and exacerbate vaccine hesitancy. Additionally, electronic media ownership has been identified as a promising avenue for improving health communication and vaccination awareness. The current study found notable levels of radio and mobile phone ownership among households, suggesting that leveraging these tools could effectively disseminate accurate health information and combat misinformation regarding vaccines (Myers M, 2008). Previous literature supports this notion, indicating that targeted health campaigns utilizing radio and mobile platforms can enhance community engagement and encourage vaccination uptake (Bangure et al., 2015). A study by Gilano G. (2024) underscores the potential of mobile health interventions in enhancing vaccination rates through text reminders and informational campaigns. The current study’s findings regarding the high ownership of radios and handsets among households in Southern Ethiopia suggest a viable avenue for targeted health communication efforts to dispel myths and convey accurate vaccine information. Accessibility remains a significant barrier to vaccination, particularly in urban settings where logistical challenges can hinder healthcare access. A study by Bangura et al. (2020) found that geographic barriers, such as distance to health facilities, significantly correlate with low immunization coverage. This is supported by other studies that consistently demonstrate that the physical distance to health services can impede immunization, with families situated farther from clinics exhibiting lower compliance rates (Oyo-Ita A, 2016). The present research echoes these findings, with many respondents reporting significant distances to health facilities, which may deter routine vaccination practices. In conclusion, addressing vaccine-preventable diseases (VPDs) requires a multifaceted approach that considers the diverse factors influencing vaccine hesitancy and non-compliance. This literature review underscores the urgent need for targeted interventions that address the socio-cultural, economic, and informational barriers to vaccination, particularly in regions like Nigeria where polio remains endemic. The findings from the study conducted by Jibrin AM (2025) highlight critical areas for intervention, including enhancing knowledge and awareness about vaccines, addressing fears and misconceptions, and leveraging the influence of community leaders and religious figures. Moreover, improving educational outreach and communication strategies is essential to counter misinformation and build trust in healthcare systems. By utilizing accessible platforms such as mobile technology and radio, health campaigns can effectively disseminate accurate information and engage communities. Additionally, addressing logistical barriers to vaccine access, particularly in urban and hard-to-reach areas, is crucial for improving vaccination rates. Ultimately, the commitment of governments, healthcare providers, and community stakeholders is vital in fostering an environment conducive to vaccination acceptance. Sustained investments in education, healthcare infrastructure, and community engagement will be necessary to achieve high vaccination coverage and protect vulnerable populations from VPDs. As we move forward, it is imperative to adopt a holistic approach that not only focuses on immunization but also on addressing the underlying determinants of health to ensure a healthier future for all.

### 1.2 Statement of the Problem

Immunization is a cornerstone of public health, serving as a critical intervention for the prevention of childhood diseases and the reduction of mortality rates. Despite the positive outcomes generated by the GAVI-supported immunization program in Bayelsa State, which included innovative strategies such as the deployment of outboard engines and Hilux vans for improved vaccine logistics, the sustainability of these gains poses a significant challenge. The program’s success was marked by increased immunization coverage, effective engagement of healthcare workers—including midwives, community health extension workers (CHEs), and mobilizers—and enhanced financial management through the use of new accounting software. However, the closure of the GAVI program in March 2025 raises pressing concerns about the future of immunization efforts in the state. The geographical and logistical challenges inherent to Bayelsa State, characterized by its difficult terrains and scattered settlements, complicate the delivery of health services. These barriers hinder access to vaccination, particularly in remote communities that remain underserved. The innovations introduced during the GAVI program have highlighted the importance of adapting strategies to local contexts. However, with the cessation of external funding and support, there is an urgent need to assess the long-term impact of these interventions and identify strategies to maintain and build upon the progress achieved. Moreover, while the GAVI-supported initiatives have shown promises, the potential for regression in immunization coverage looms large without a robust, sustainable plan in place. The reliance on external funding mechanisms poses a risk to the continuity of vaccination services, particularly as local health systems may lack the financial resources and infrastructure to effectively respond to the ongoing immunization needs of the population. The challenges of vaccine hesitancy, caregiver misconceptions, and a lack of awareness about the importance of immunization further exacerbate the situation, necessitating targeted efforts to address these issues in the post-GAVI landscape. In this context, the imperative to study the outcomes of the GAVI program is paramount. There is a critical need to evaluate the effectiveness of the strategies employed, understand the barriers that remain, and identify facilitators that could support the sustained delivery of immunization services. This evaluation must not only focus on quantitative metrics of coverage but also delve into the qualitative aspects of caregiver knowledge, attitudes, and practices regarding vaccination. By leveraging the insights gained from the GAVI experience, stakeholders can develop a comprehensive and sustainable immunization plan that takes into account the current realities of limited financial resources and complex geographical challenges. The future of immunization in Bayelsa State hinges on the ability to translate the successes of the GAVI-supported program into a resilient and self-sustaining health system capable of addressing the immunization needs of all children, particularly those in hard-to-reach areas. Without a strategic approach that incorporates lessons learned, community engagement, and innovative solutions to logistical challenges, the achievements made during the GAVI program risk being undermined, leaving vulnerable populations at increased risk for vaccine-preventable diseases. Therefore, this study aims to assess the outcomes of the GAVI-supported immunization initiatives and propose actionable strategies for sustaining high immunization coverage in Bayelsa State amidst the pressing challenges that lie ahead.

### 1.3 Research Questions

a. What is the current immunization coverage among children aged 0-59 months in Bayelsa State following the GAVI-funded immunization program?
b. What proportion of children aged 0-59 months possess their immunization cards?
c. What is the prevalence of non-compliance with recommended immunization schedules among children aged 0-59 months, and what are the common reasons cited for non-compliance?
d. How does the current immunization coverage compare with the baseline coverage of 61.12% prior to the implementation of the GAVI-funded program?
e. What socio-demographic factors (e.g., age, gender, caregiver education, socioeconomic status, location) are associated with immunization coverage, card retention, and non-compliance?
f. What are the levels of knowledge, attitudes, and practices of caregivers regarding childhood immunization in the context of the GAVI-funded program?
g. What strategies from the GAVI-supported interventions can be leveraged to sustain and improve immunization coverage in the future?

### 1.4 Research Objectives

#### General Objective

To assess the impact of GAVI-supported immunization interventions on childhood vaccine coverage, card retention, and compliance in Bayelsa State.

#### Specific Objectives

a. To determine the current immunization coverage among children aged 0-59 months in Bayelsa State post-GAVI support.
b. To evaluate the level of vaccine card retention among children in the target age group.
c. To assess the compliance of caregivers with the recommended immunization schedule.
d. To identify the key barriers and facilitators influencing immunization uptake in the context of GAVI-supported interventions.
e. To provide evidence-based recommendations for sustaining and improving immunization coverage in Bayelsa State.

### 1.5 Significance of the Study

The GAVI-supported immunization program in Bayelsa State introduced a range of innovative strategies and technologies that significantly enhanced vaccine delivery in a challenging environment. The deployment of inboard engines and Hilux vans revolutionized vaccine logistics, enabling health teams to reach remote, hard-to-reach settlements and underserved communities that were previously difficult to reach. Additionally, new approaches to engaging and deploying healthcare workers, along with the adoption of improved financial management software, contributed to the overall success of the program. These interventions marked a notable milestone in strengthening immunization services, reducing the number of zero-dose children, and improving coverage rates. However, as the program’s direct support diminishes, there is an urgent need to sustain and build upon these gains. This study aims to evaluate the outcomes of the GAVI-supported interventions to determine what strategies were most effective and how they can be leveraged for long-term sustainability. Given the limited financial resources, diverse geographical challenges, and logistical constraints, it is crucial to develop a sustainable immunization plan that ensures continued coverage, card retention, and compliance among children aged 0-59 months. The findings of this study will provide valuable insights to policymakers, health program managers, and stakeholders, guiding strategic decision-making and resource allocation to maintain and expand the successes achieved during the GAVI-supported period. Furthermore, understanding the outcomes of these initiatives will help identify best practices and innovative strategies that can be adapted and scaled up in other similar contexts facing geographical, security, and resource-related challenges. By systematically assessing the effectiveness of these interventions, the study will contribute to evidence-based planning and policy formulation aimed at maintaining high immunization coverage beyond external support. This is vital in ensuring that the gains achieved are not only sustained but also translated into resilient and self-reliant immunization systems capable of addressing future challenges. Ultimately, the insights generated will aid in designing pragmatic and sustainable frameworks that utilize limited resources efficiently, address geographic and security constraints, and guarantee that every child in Bayelsa State receives timely and complete immunizations, thereby safeguarding their health and contributing to broader public health goals.

### 1.6 Scope and Limitations of the Study

This study is geographically focused on Bayelsa State, specifically targeting selected communities within the state that represent both urban and rural settings. The study population comprises caregivers of children aged 0-59 months, with data collected on immunization status, card retention, caregiver knowledge, attitudes, and practices related to childhood immunization.

#### 1.6.1 Scope of study

The scope includes:

a. Assessing current immunization coverage using household surveys.
b. Verifying immunization status through immunization cards and caregiver recall.
c. Exploring socio-demographic factors influencing immunization uptake.
d. Investigating reasons for non-compliance and barriers to vaccination.
e. Evaluating caregiver knowledge and attitudes towards immunization.
f. Analysing the retention and use of immunization cards as a record-keeping tool.

#### 1.6.2 Limitations of the Study

While the study aims to provide comprehensive insights, several limitations should be acknowledged:

a. Recall Bias: In cases where immunization cards are unavailable, data rely on caregiver recall, which may be inaccurate or biased, potentially affecting the validity of immunization coverage estimates.
b. Selection Bias: The multistage sampling approach, though designed to be representative, may still lead to selection bias if some households are missed or refuse participation, potentially affecting the generalizability of findings.
c. Cross-Sectional Design: As a snapshot in time, the study cannot establish causal relationships or assess changes in immunization patterns over time. Longitudinal studies would be needed for such analyses.
d. Limited Geographical Coverage: The study covers selected communities within Bayelsa State and may not fully capture the diversity of all communities, especially more remote or hard-to-reach areas, limiting the generalizability to the entire state.
e. Resource Constraints: Limitations in funding, personnel, and logistical support may restrict the sample size or depth of data collection, potentially impacting the comprehensiveness of the findings.
f. Security issues: Security issues in certain areas could restrict access to some communities, thereby limiting comprehensive data collection.
g. Potential Reporting Bias: Caregivers might overreport immunization status due to social desirability or perceived expectations from interviewers.
h. Temporal Limitations: Data collected at a single point in time do not account for seasonal variations or ongoing program changes that may influence immunization coverage.

### 2.0 Methodology

### 2.1 Study Design

This study employed a descriptive cross-sectional design to assess the immunization coverage, card retention, and factors associated with immunization non-compliance among children aged 0-59 months in Bayelsa State. Data were collected at a single point in time from a representative sample of caregivers, allowing for the evaluation of the current immunization status and identifying potential correlates within the population. The cross-sectional nature allows for an accurate assessment of immunization status, caregiver knowledge, and barriers at one point in time, providing a baseline for program evaluation. It is resource-efficient, requiring less time and cost compared to longitudinal studies, making it suitable for community-based surveys. The design facilitates examination of relationships between immunization coverage and various socio-demographic, behavioral, and systemic factors. The study will involve structured interviews with caregivers and verification of immunization cards within selected communities.

### 2.2 Study Area

Bayelsa State is located in the southern part of Nigeria, in the Niger-Delta region. It is bordered by Rivers State to the West and Delta State to the East with a long span of Atlantic Ocean at the south. The capital city is Yenagoa. Bayelsa has a population of about 2,537,400 with a landscape area of 9,391 km^2^ (NPC, 2022). Demographic data for Bayelsa State indicates that most of the population belongs to the Ijaw ethnic group, which is the dominant ethnic group in the state. Other minority ethnic groups include the Ogbia, Nembe, and Epie-Atissa. The main languages spoken in Bayelsa State are Ijaw, Epie-Attisa, Isoko, Urhobo and English. Bayelsa State has a predominantly Christian population, with Christianity being the major religion practiced in the state. However, there are also adherents of other religions, including traditional Africans religions and Islam. The economy of Bayelsa State is predominantly petroleum resources, as the state is in the oil-rich Niger Delta region. Bayelsa has one of the largest crude oil and natural gas deposits in Nigeria, with the Oloibiri Oilfield being the site of the country’s first oil discovery. Other mineral raw materials found in the state include salt, agro raw materials include cassava, plantain, rice, and fish.

### 2.3 Study Population

The study was conducted in Bayelsa State, located in the South-South geopolitical zone of Nigeria. The state comprises both urban and rural communities, with diverse socio-economic backgrounds. The health infrastructure includes primary health care centres, clinics, and outreach services supported by the GAVI immunization program. The target population comprised caregivers of children aged 0-59 months residing in Bayelsa State. Children who had been residents of the communities for at least six months prior to the survey were eligible. Caregivers who declined consent or were unavailable after repeated visits were excluded.

### 2.4 Sample Size Determination

The immunization coverage in DHIS2. (2025), before the commencement of the GAVI-funded immunization services in the fourth quarter 2021 was 61.12%. To calculate the sample size needed for this study, the formula for sample size determination using proportions as stated:

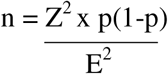

Where:

n = required sample size

Z = Z-value (the number of standard deviations from the mean for a given confidence level) p = estimated proportion (immunization coverage)

E = margin of error (the difference you are willing to accept between the sample proportion and the true population proportion)

Given:

p = 0.6112 (or 61.12%) immunization coverage Z = 1.96 (confidence interval of 95%

E = 0.05 (5%)

Calculation:

Using the assumed margin of error of 0.05, we can calculate the sample size:

Substitute the values into the formula:

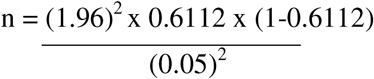

Calculate:

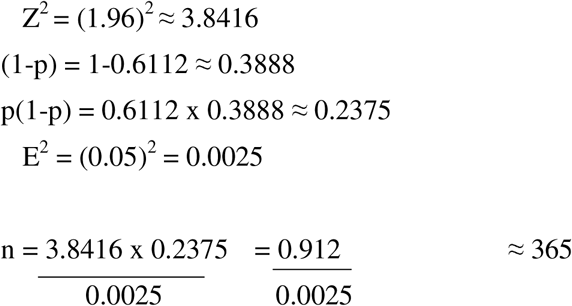

Additional samples = 365 + 15% ≈ 420

In order to make up for non-responses, contingency issues, dropouts an additional sample of 15% were added to the sample size resulting in a final sample size of approximately 420 children. A larger sample will reduce the margin of error, thereby increase the precision of the estimated proportion and enhance the overall accuracy of the study outcomes.

### 2.5 Sampling Technique

To ensure the collection of representative and reliable data on childhood immunization and noncompliance among children of aged 0-59 months in South-South Bayelsa, a multistage sampling approach complemented by stratified sampling was employed. This methodology was selected to effectively capture the diverse community settings and demographic characteristics across the state, while ensuring that only children within the specified age range were included. Given the geographical diversity and the various community settings involved in vaccination campaigns, a multistage sampling process was adopted. Stage 1 involves the selection of eight local counties called the Local Government Areas (LGA). This initial step ensured broad geographic coverage and representation of different community contexts. The second stage involves the identification of key settings within the LGAs. In each selected LGA, specific sites where vaccination campaigns are conducted were identified. These included households, churches, mosques, marketplaces, waterfronts, public motor parks, and border areas. These sites were chosen based on their known roles in vaccination outreach activities. The third stage involves the selection of households and children. From each identified settings, households with children aged 0-59 months were listed. Using systematic or simple random sampling, households containing eligible children were selected. Within each household, the target child was identified, and data was collected based on parental or caregiver consent. This consent was given verbally. In addition to household sampling, purposive sampling was used to select key informants such as religious leaders, community health workers, and vaccination team members. These individuals provide qualitative insights into community perceptions, practices, and challenges related to childhood immunization.

### 2.6 Selection Criteria

*Inclusion Criteria*: Children aged 0-59 months residing within the selected communities during the study period. Caregivers or parents of eligible children who provide informed consent.

*Exclusion Criteria*: Children outside the age range of 0-59 months. Children whose caregivers decline participation or are unavailable after repeated contact attempts.

### 2.7 Method of Data Collection

Data were collected using a structured, interviewer-administered questionnaire developed based on WHO standards and previous surveys with Google form. The questionnaire was pre-tested in a neighboring community not included in the main study. It comprised sections on socio-demographic characteristics of caregivers and children, immunization status and history, immunization card possession and retention, reasons for missed or delayed immunizations, caregivers’ knowledge and attitudes towards immunization, access to and utilization of immunization services. Additionally, immunization cards were examined to verify vaccination history. Trained enumerators conducted face-to-face interviews with caregivers after obtaining informed consent verbally. The immunization status was verified through immunization cards whenever available; otherwise, caregiver recall was used.

### 2.8 Pre-testing

***Validity test*:** The questionnaire and other instruments of measurement were subjected to testing in a community not selected for sampling. A statistically significant value of p<0.05 and confidence interval of 95% was used to test the validity of study.

***Reliability test*:** The study was repeated in different selected communities using the same questionnaire and tools of measurement and comparative analysis was done. Results from these separate studies showed consistency with the results which established the reliability of the questionnaire and tools of measurement. The study showed that when the questionnaires and tools of measurement were subjected to use at various location and climatic condition it had proved to measure the same outcomes with minimal variation.

### 2.9 Data Management and Analysis

Data were coded, checked for completeness and consistency, and entered into Microsoft Excel and SPSS version 23 for analysis. Descriptive statistics such as frequencies, percentages, means, and standard deviations were used to summarize the data. Inferential statistics was tested using one-sample proportion z-test to compare current immunization coverage with the baseline. Chi-square tests were used to examine associations between categorical variables (e.g., socio-demographics and immunization compliance). A p-value of less than 0.05 was considered statistically significant.

### 2.10 Timeline for the Study

***Research Planning and Proposal Development (week 1 and 2):*** - The study commenced in June 2025, with the initial two weeks dedicated to comprehensive planning. During Weeks 1 and 2, the research team defined clear objectives, develop the detailed research protocol, design data collection tools such as questionnaires, and determined the sampling strategies and logistical plans. This phase ensures that all methodological, operational, and ethical considerations were thoroughly addressed to facilitate smooth implementation.

***Ethical Approval (week 3 and 4)***: - In Weeks 3 and 4 of June 2025, the team prepared and submitted the research proposal to Ethics Committee, Bayelsa State Primary Health Care Board for ethical clearance. The submission included the study protocol, consent forms, data management plans, and other supporting documents. The team actively followed up to obtain official approval by the end of Week 4, ensuring that all ethical standards were met before proceeding to data collection.

***Data Collection Preparation***: - The first two weeks of July 2025 focused on preparing for data collection. This included recruiting and training data collectors and supervisors on data collection procedures, ethical considerations, questionnaire administration, and data quality assurance. During Week 2 of July, a pilot test of questionnaires was conducted in a small number of households to identify ambiguities or logistical issues, allowing for necessary adjustments. Finalization of data collection tools, logistical arrangements, and fieldwork schedules were completed by mid-July, ensuring readiness.

***Data Collection***: - Data collection took place for two weeks, from mid-July to end of July 2025. Trained enumerators visited selected households within sampled communities to gather information on children aged 0-59 months and their caregivers, following the approved protocols. Supervisors were deployed to oversee daily activities, conduct regular quality checks, and address challenges promptly to ensure completeness, accuracy, and reliability of the data collected throughout this period.

***Report Writing and Publication:*** - In the first weeks of August 2025, the team cleaned, analysed, and interpret the collected data. Based on preliminary findings, a comprehensive draft was initiated, including findings and recommendations. The second week saw the proof reading of the draft, editing, and finalization. The finalized draft was prepared as manuscript for submission for publication in peer-reviewed journals.

***Dissemination and Policy Engagement***: - During the final two weeks of September 2025, the study findings will be disseminated through stakeholder workshops, policy dialogues, and presentations to government agencies, donors, and partner organizations after journal publication. These activities aim to promote evidence-based decision-making and inform policy formulation.

### 2.11 Ethical Consideration

***Institutional Consent:*** - Prior to initiating the study, formal approval will be sought from Ethics Committee, Bayelsa State Primary Health Care Board to ensure adherence to ethical standards concerning research conduct, participant safety, and data confidentiality. A detailed research protocol outlining the study objectives, methodology, sampling procedures, data management, and measures to protect participants’ rights was submitted for review. Only upon receipt of the official approval from the Ethics Committee was data collection commenced. This process ensures that the study complies with national and institutional ethical guidelines, safeguarding the dignity, rights, and welfare of all sampled households and individuals involved.

***Community Consent:*** - Since households are located within communities, engaging with community leaders and local authorities was essential to obtaining community-level consent and support which was done verbally. Prior to household sampling, community meetings and consultations were conducted to inform community elders, leaders, and relevant stakeholders about the purpose of the study, its potential benefits, and the measures in place to protect participants’ rights. Their approval and support were sought to facilitate access to households and ensure culturally appropriate conduct. Gaining community consent fosters trust, promotes cooperation, and respects local customs, which was vital for ethical and effective data collection.

***Individual Consent***: - Participation of household members, particularly caregivers of children aged 0-59 months, was voluntary. For each household selected, verbal consent was obtained from the caregiver or parent/guardian before any data collection activities began. Participants were provided with clear and comprehensible information about the study’s purpose, procedures, potential risks and benefits, and their right to refuse or withdraw at any time without any penalty. Consent was documented either in writing or verbally (if literacy is limited), ensuring respect for individual autonomy. Confidentiality and data privacy were strictly maintained, and all personal information were anonymized during analysis and reporting.

### 3.0 Results

Table 1 presents the socio-demographic characteristics of caregivers and children involved in the study. Among the children, there is a slight male predominance, with 51% being male and 49% female. The age distribution reveals that a significant portion of the children, 44.3%, are under one year old, highlighting the vulnerability of this age group to vaccine-preventable diseases. The majority of caregivers are male heads of households (72.6%), reflecting traditional gender roles in decision-making within families. The age of household heads shows that 47.9% fall within the 31-40 years range, indicating that many caregivers are likely in their prime parenting years. In terms of education, 54.8% of household heads have attained secondary education, while 30% possess tertiary education, suggesting a generally educated caregiver population. The religious affiliation of caregivers is predominantly Christian, accounting for 90.2% of the respondents. The occupation data indicates that most household heads (57.6%) are self-employed, which may affect their economic stability and access to healthcare services. Lastly, the majority of the population resides in semi-urban areas (80.7%), which could present unique challenges in accessing health facilities and immunization services compared to urban or rural settings. Overall, these characteristics provide valuable context for understanding the factors influencing immunization coverage and compliance in the state.

**Table 1:** Socio-Demographic Characteristics of Caregivers and Children

| Variables | Frequency (n=420) | Percentage (%) |
| --- | --- | --- |
| **Age of child** |  |  |
| *Less than 1 year* | 186 | 44.3% |
| *1-2 years* | 145 | 34.5% |
| *3-4 years* | 77 | 18.3% |
| *5 years* | 12 | 2.9% |
| **Gender of child** | Frequency (n=420) | Percentage (%) |
| *Male* | 214 | 51.0% |
| *Female* | 206 | 49.0% |
| **Household Head** | Frequency (n=420) | Percentage (%) |
| *Male* | 305 | 72.6% |
| *Female* | 115 | 27.4% |
| **Age of Household Head** | Frequency (n=420) | Percentage (%) |
| *18-30 years* | 137 | 32.6% |
| *31-40 years* | 201 | 47.9% |
| *41-50 years* | 75 | 17.9% |
| *51 years and above* | 7 | 1.7% |
| **Religion** | Frequency (n=420) | Percentage (%) |
| *Christianity* | 379 | 90.2% |
| *Muslim* | 41 | 9.8% |
| **Educational level of household Head** | Frequency (n=420) | Percentage (%) |
| *No formal education* | 26 | 6.2% |
| *Primary education* | 38 | 9.0% |
| *Secondary education* | 230 | 54.8% |
| *Tertiary education* | 126 | 30.0% |
| **Occupation of Household Head** | Frequency (n=420) | Percentage (%) |
| *Employed* | 104 | 24.8% |
| *Unemployed* | 58 | 13.8% |
| *Self-employed* | 242 | 57.6% |
| *Student* | 9 | 2.1% |
| *Others* | 7 | 1.7% |
| **Types of Settlement** | Frequency (n=420) | Percentage (%) |
| *Urban* | 44 | 10.5% |
| *Semi-urban* | 339 | 80.7% |
| *Rural* | 37 | 8.8% |

The vaccination uptake data presented for the months of October to December in 2021 and 2024 shows a significant increase in the number of vaccinations administered across all antigens. There is a clear upward trend in vaccination uptake from 2021 to 2024. This trend reflects an overall improvement in vaccination efforts and possibly heightened awareness or accessibility to vaccines over the three-year period. Starting with the BCG vaccine, the uptake rose from 6,447 doses in 2021 to 16,407 in 2024. This is more than twofold increase and suggests an enhanced focus on tuberculosis prevention among infants, possibly due to public health campaigns or improved healthcare infrastructure. Similarly, the OPV-3 (Oral Polio Vaccine) saw a substantial surge from 7,962 doses to 20,324. This increase is particularly encouraging as it indicates a concerted effort to eradicate polio and ensure that a larger proportion of children are protected against this disease. The Penta-3 vaccine, which protects against multiple diseases (diphtheria, tetanus, pertussis, hepatitis B, and Haemophilus influenzae type b), displayed a similar growth pattern, with uptake climbing from 7,964 in 2021 to 20,317 in 2024. This consistent rise underscores the importance of comprehensive immunization strategies in safeguarding child health. The Measles vaccination also experienced a notable increase, with doses administered rising from 6,947 to 11,361. While this growth was a positive one, the increase when compared to other vaccines, suggest that there may still be challenges in reaching certain populations or addressing vaccine hesitancy. Lastly, the Yellow Fever vaccine uptake jumped from 7,627 to 17,892 doses. This increase could be attributed to improved public health initiatives and perhaps rising awareness about the importance of vaccination in preventing outbreaks of yellow fever, particularly in endemic areas. Overall, the total vaccination uptake across all antigens escalated from 36,947 in 2021 to 86,301 in 2024, indicating a robust and effective vaccination program over these years. This upward trajectory highlights the success of health policies and interventions aimed at increasing immunization coverage, ultimately contributing to better public health outcomes. However, continuous efforts are needed to maintain and further improve these numbers, particularly for vaccines like measles, where the growth is not as pronounced.

Table 2 presents the immunization coverage rates for various vaccines among children aged 0-59 months in Bayelsa State. The data indicates that coverage rates for different vaccines varied significantly. The Bacillus Calmette-Guérin (BCG) vaccine and the Oral Polio Vaccine (OPV) showed the highest coverage, with rates of 93.6% and 94.3%, respectively. In contrast, the coverage for the measles vaccine was notably lower at 56.0%, while yellow fever coverage was slightly higher at 54.5%. The coverage for the Pentavalent vaccine was moderate at 82.6%. These findings suggest a positive trend in immunization uptake for certain vaccines, particularly BCG and OPV, which are typically administered at birth and in the early months of life. However, the lower coverage rates for the measles and yellow fever vaccines raise significant concerns, as these are critical immunizations for preventing severe diseases. The discrepancy in coverage rates highlights challenges in achieving high compliance for later vaccines in the immunization schedule, indicating a need for enhanced community education and outreach to address misconceptions and barriers that hinder vaccination uptake. Overall, while the data reflects successful outreach strategies for certain vaccines, it underscores the necessity for targeted interventions to improve coverage for all vaccines in the immunization schedule.

**Table 2:** Immunization Coverage Rates by Antigen Among Children Aged 0-59 Months

| Antigen | Frequency (n=420) | Coverage |
| --- | --- | --- |
| BCG | 393 | 93.6% |
| OPV | 396 | 94.3% |
| Penta vaccine | 347 | 82.6% |
| Measles | 235 | 56.0% |
| Yellow fever | 229 | 54.5% |

*Source: sourced from household survey data*

Table 3 presents the overall immunization coverage among children aged 0-59 months in Bayelsa State. The results indicate that a significant percentage of children, specifically 95.5%, are fully immunized, with only 4.5% classified as having zero doses. This high level of overall immunization coverage is promising and suggests that the GAVI-supported immunization initiatives have had a favourable impact on vaccination rates in the state. When compared with the baseline immunization coverage of 61.12% reported prior to the GAVI intervention, the current figures reflect a substantial improvement in vaccine uptake.

**Table 3:** Overall Immunization Coverage Among Children Aged 0-59 Months

| Status | Frequency (n=420) | Coverage |
| --- | --- | --- |
| Total immunization | 401 | 95.5% |
| Zero dose | 19 | 4.5% |

Table 4 reveals the perceptions of caregivers regarding vaccination among children aged 0-59 months in Bayelsa State. The data indicates that a significant portion of caregivers holds varying beliefs about vaccines, which can influence their commitment to immunization practices. Specifically, 19.1% of caregivers expressed the belief that vaccines are not safe, while a larger proportion, 40.4%, reported concerns regarding potential side effects of vaccines. Additionally, 17.0% of caregivers deemed vaccines unnecessary, and 23.4% indicated that they had religious or philosophical beliefs against vaccination. These findings highlight a critical aspect of vaccine acceptance: even among caregivers who have accepted vaccination for their children, lingering doubts and misconceptions about vaccine safety and necessity persist.

**Table 4:** Caregivers’ Perception of Vaccine Safety and Necessity

| Perception | Frequency (n=47) | Percentage (%) |
| --- | --- | --- |
| **I believe vaccines are not safe** | 9 | 19.1% |
| **I have concerns about vaccine side effects** | 19 | 40.4% |
| **I think vaccines are unnecessary** | 8 | 17.0% |
| **I have religious or philosophical beliefs against vaccination** | 11 | 23.4% |

Table 5 presents the various reasons cited by caregivers for non-compliance with vaccination schedules among children aged 0-59 months in Bayelsa State. The data reveals several key trends that shed light on the barriers to timely immunization. The most frequently reported reason for non-compliance, cited by 34.6% of respondents, was “I forgot my next visit according to the schedule.” This highlights a significant gap in awareness or engagement with the immunization schedule, suggesting that caregivers may lack effective reminders or support systems to ensure timely vaccinations. Other notable reasons included “concerns about vaccine side effects” (21.2%), which reflects prevalent fears stemming from misinformation or lack of education about vaccines. Additionally, 14.0% of caregivers cited “cultural beliefs” as a reason for non-compliance. Cultural narratives and traditional beliefs can significantly shape health behaviours, leading to skepticism toward modern medical practices, including vaccination. Furthermore, 7.7% of respondents indicated, “I did not receive adequate information about vaccination,” reinforcing the need for targeted educational outreach. Inadequate dissemination of information can perpetuate misconceptions, leading to increased vaccine hesitancy and non-compliance. Other reasons included “I did not have access to healthcare services” (3.8%) and “religious beliefs” (5.8%), illustrating systemic barriers to vaccination. These findings underscore the necessity for comprehensive educational initiatives and community engagement strategies that address both informational needs and cultural sensitivities.

**Table 5:**
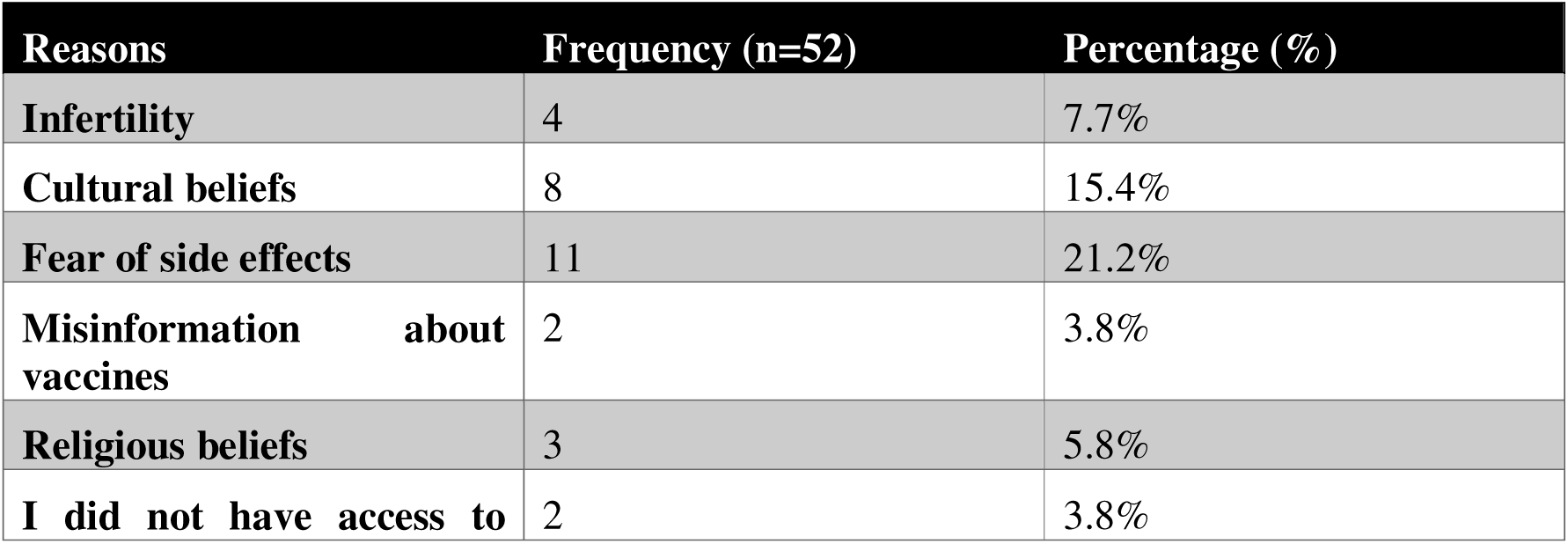
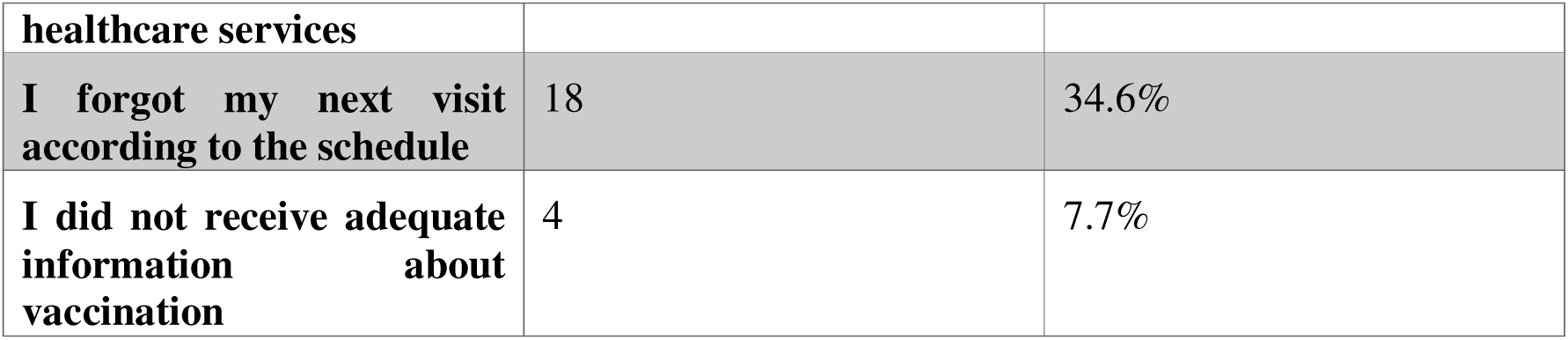
Reasons for Non-Compliance with Vaccination Schedules Among Caregivers

Table 6 highlights the significant relationship between the distance to health facilities and accessibility to vaccination services among children aged 0-59 months in Bayelsa State. The analysis reveals a chi-square value of 10.84, with a p-value of 0.001, indicating a statistically significant association. The data shows that households located less than 1 km from health facilities have higher vaccination participation rates compared to those living farther away. This finding underscores the critical role that geographic proximity plays in influencing vaccination compliance. Families residing within closer distances are more likely to access vaccination services regularly, whereas those living further away face logistical challenges that can hinder their ability to seek timely immunizations. The results emphasize the need for targeted interventions to improve access to vaccination services, particularly for families living in more remote areas, in order to enhance overall immunization coverage and ensure that children are protected against vaccine-preventable diseases.

**Table 6:**
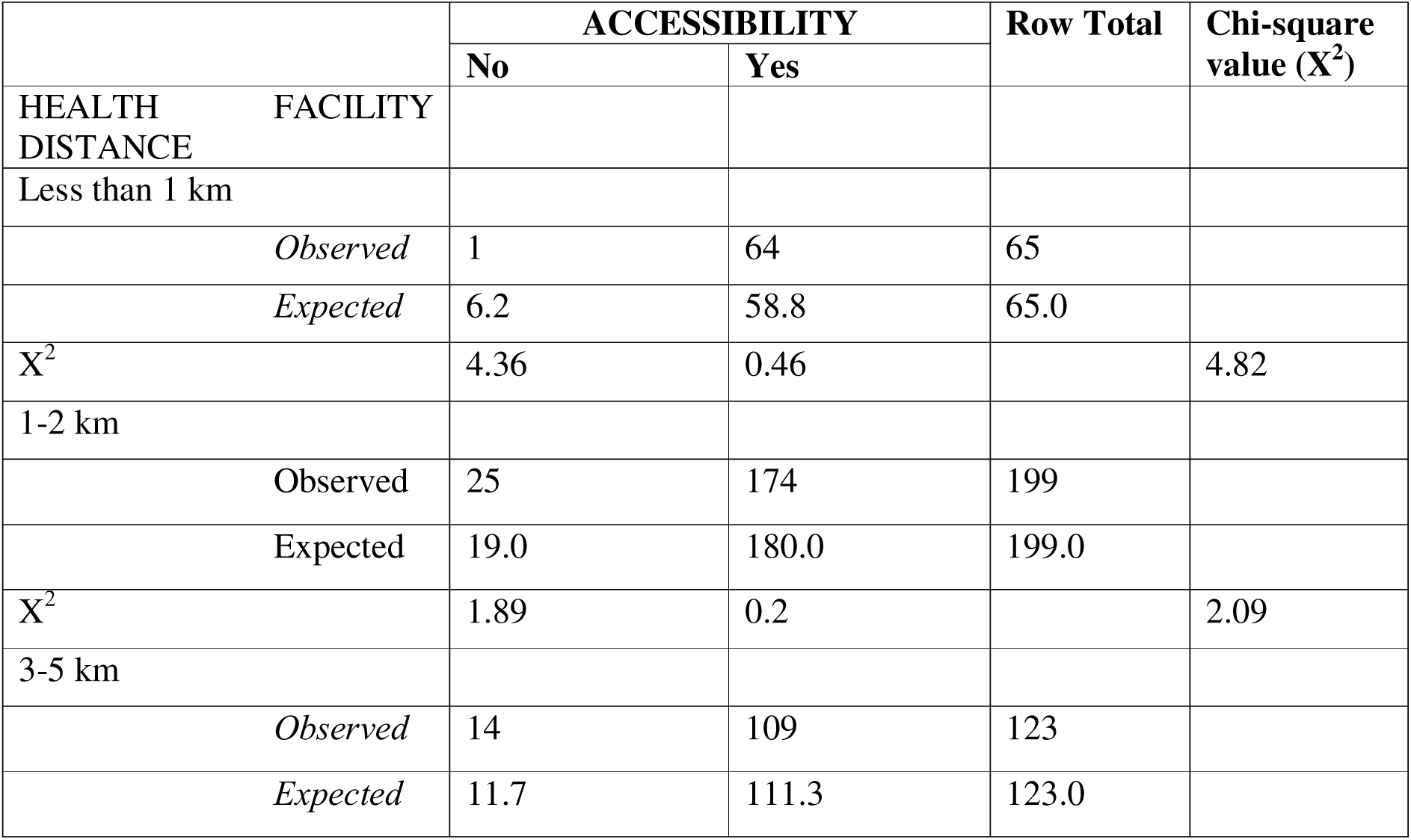
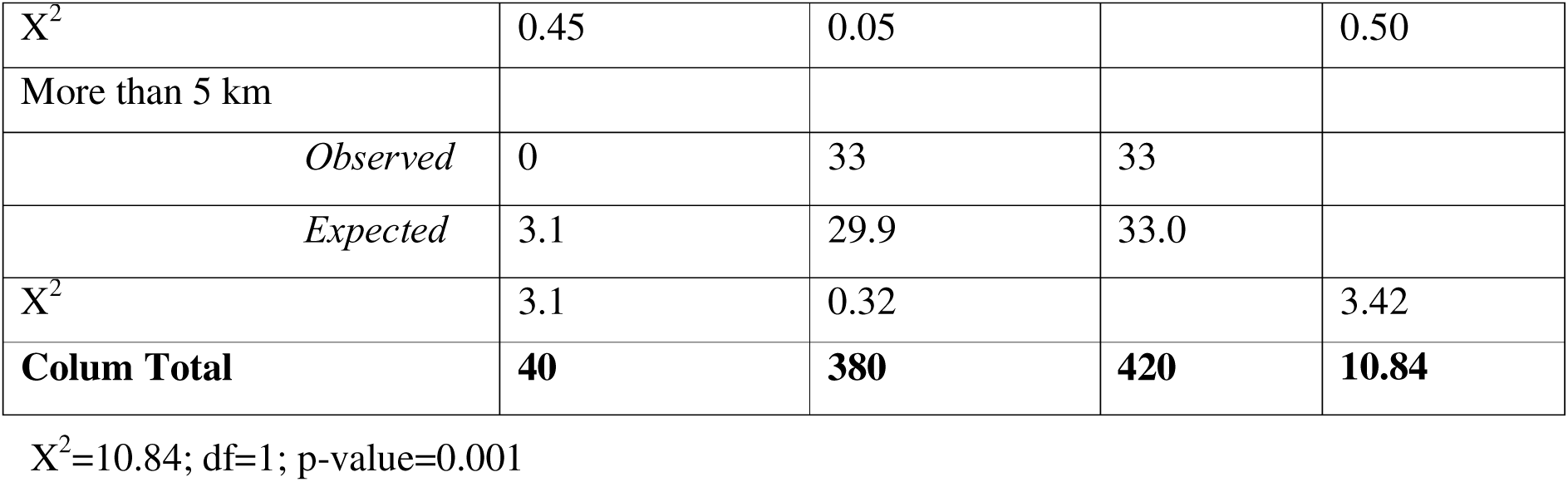
Relationship Between Distance to Health Facilities and Vaccination Accessibility

Table 7 presents the analysis of the relationship between the educational level of household heads and the retention of immunization cards among children aged 0-59 months in Bayelsa State. The results indicate a significant association, with a Chi-square value of 17.92 and a p-value of less than 0.00002. This finding suggests that the level of education among caregivers plays a crucial role in determining whether immunization cards are retained. Specifically, caregivers with higher educational attainment demonstrated significantly better card retention rates compared to those with lower educational levels. For instance, households led by individuals with tertiary education exhibited the highest retention rates, while those with no formal education showed the lowest retention. This pattern highlights the importance of educational outreach in improving not only the understanding of vaccination processes but also the importance of maintaining accurate health records. Overall, this data emphasizes the need for targeted educational interventions that can enhance card retention and, consequently, improve vaccination compliance among children in the community.

**Table 7:** Association Between Educational Level of Household Heads and Immunization Card Retention

| | Card Retention | | | Chi-square value ($X^2$) |
| --- | --- | --- | --- | --- |
|  | No | Yes | Row Total |  |
| **Educational Level** |  |  |  |  |
| **No Formal Education** |  |  |  |  |
| *Observed* | 7 | 19 | 26 |  |
| *Expected* | 2.5 | 23.5 |  |  |
| $X^2$ | 7.84 | 0.85 | | 8.69 |
| **Primary Education** |  |  |  |  |
| *Observed* | 3 | 35 | 38 |  |
| *Expected* | 3.71 | 34.29 |  |  |
| $X^2$ | 0.00 | 0.01 | | 0.01 |
| **Secondary Education** |  |  |  |  |
| *Observed* | 12 | 217 | 229 |  |
| *Expected* | 22.35 | 206.65 |  |  |
| $X^2$ | 4.80 | 0.52 | | 5.32 |

| **Tertiary Education** |  |  |  |  |
| --- | --- | --- | --- | --- |
| *Observed* | 19 | 108 | 127 |  |
| *Expected* | 12.40 | 114.60 |  |  |
| $X^2$ | 3.52 | 0.380 | | 3.90 |
| Column Total | **41** | **379** | **420** | **17.92** |

$X^2=17.92$ ; $df=1$ ; $p\text{-value}=0.00002$

The findings from Table 8 underscore the critical role that various sources of vaccination information play in influencing caregivers’ participation in immunization programs. The data reveals that the majority of caregivers, 204 respondents, reported that information from family and friends significantly impacted their decision to vaccinate their children. This strong reliance on personal networks highlights the importance of social connections and trust in shaping health behaviours, particularly in communities where traditional beliefs and skepticism around vaccines may exist. Moreover, healthcare workers emerged as another vital source of information, with 374 caregivers indicating that they turn to medical professionals for guidance on vaccinations. This underscores the essential role that healthcare providers play in educating caregivers about the safety and necessity of vaccines. The trust that caregivers place in healthcare professionals can be leveraged to enhance vaccination rates, suggesting that strengthening the communication skills of these providers could lead to improved health outcomes. Conversely, the data indicates that reliance on other information sources such as radio (189), social media (100), town announcements (18), and church communications (7) was notably lower. This disparity suggests that while these channels can be useful, they may not be as effective in reaching caregivers or influencing their decisions as personal relationships and direct professional advice. The chi-square value of 5.04 indicates a statistically significant relationship between the sources of vaccination information and participation in immunization programs. This finding reinforces the notion that caregivers are more likely to engage in immunization programs when they receive information from trusted sources, particularly those within their immediate social circle and healthcare professionals. In light of these insights, it is crucial to develop targeted communication strategies that harness the influence of family and friends, as well as healthcare providers, to foster a supportive environment for vaccination. Public health campaigns could benefit from involving these trusted figures in dissemination efforts, thereby enhancing the credibility of vaccination messages and increasing overall immunization uptake. By leveraging existing social networks and building strong relationships with healthcare providers, stakeholders can effectively address vaccine hesitancy and improve vaccination rates within communities.

**Table 8:** Influence of Vaccination Information Sources on Participation in Immunization Programs

|  | **Vaccination**<br>**(n=420)** | **participation** | **Total** | **Chi-square**<br>**value ($X^2$)** |
| --- | --- | --- | --- | --- |
|  | No | Yes |  |  |
| **Vaccination information** |  |  |  |  |
| **Family and friends** |  |  |  |  |
| *Observed* | 23 | 204 | 227 |  |
| *Expected* | 21.53 | 205.47 |  |  |
| $X^2$ | 0.10 | 0.01 | | **0.11** |
| **Radio** |  |  |  |  |
| *Observed* | 23 | 189 | 212 |  |
| *Expected* | 20.11 | 191.89 |  |  |
| $X^2$ | 0.42 | 0.04 | | **0.46** |
| **Health Care Worker** |  |  |  |  |
| *Observed* | 31 | 374 | 405 |  |
| *Expected* | 38.41 | 366.59 |  |  |
| $X^2$ | 1.43 | 0.15 | | **1.58** |
| **Social Media** |  |  |  |  |
| *Observed* | 12 | 100 | 112 |  |
| *Expected* | 10.62 | 101.38 |  |  |
| **X<sup>2</sup>** | 0.18 | 0.019 |  | **0.20** |
| **Town Announcement** |  |  |  |  |
| *Observed* | 4 | 18 | 22 |  |
| *Expected* | 2.09 | 19.91 |  |  |
| **X<sup>2</sup>** | 1.75 | 0.18 |  | **1.94** |
| **Church** |  |  |  |  |
| *Observed* | 0 | 7 | 7 |  |
| *Expected* | 0.66 | 6.34 |  |  |
| **X<sup>2</sup>** | 0.66 | 0.07 |  | **0.73** |
| **Television** |  |  |  |  |
| *Observed* | 10 | 91 | 101 |  |
| *Expected* | 9.58 | 91.42 |  |  |
| **X<sup>2</sup>** | 0.02 | 0.002 |  | **0.02** |
| **Column Total** | **103** | **983** | **1086** | **5.04** |

$X^2=5.04$ ; $df=1$ ; $p\text{-value}= 0.025$

To conduct a z-test for the immunization coverage in Bayelsa State, we will compare the current immunization coverage with the baseline coverage prior to the GAVI intervention.

1. Calculate for the standard error

SE = √p(1-p)/n = √0.955(1-0.955)/420 = √ (0.955×0.04)/420 = √0.042975/420 = √0.00010236

≈ 0.01012

2. Calculate for the z-value

Z = (p-p0)/SE = (0.955-0.6112)/0.01012 = (0.3438)/0.01012 ≈ 33.98

Table 9 presents the results of a z-test comparing the current immunization coverage of 95.5% with the baseline coverage of 61.12%. The standard error was calculated to be approximately 0.01012. Using this standard error, the z-value was computed to be approximately 33.98. This value is significantly greater than the critical z-value of ± 1.96 for a 95% confidence interval. Consequently, we reject the null hypothesis, which states that there is no difference in immunization coverage before and after the GAVI intervention. This indicates a statistically significant increase in immunization coverage in Bayelsa State following the implementation of the GAVI-supported immunization program. The results highlight the substantial positive impact of these initiatives on vaccination rates among children aged 0-59 months in the state.

**Table 9:** Z-test results for Comparison of Current Immunization Coverage with Baseline Coverage

| Parameter | Value |
| --- | --- |
| Baseline immunization coverage (p<sub>0</sub>) | 0.6112 (61.12%) |
| Current immunization coverage (p) | 0.955 (95.5%) |
| Sample size (n) | 420 |
| Critical z-value (α=0.05) | ±1.96 |

### 4.0 Discussion

The findings from this study provide significant insights into the immunization coverage, card retention, and factors affecting vaccine compliance among children aged 0-59 months in Bayelsa State, Nigeria. Each aspect of the results highlights critical areas for public health intervention and underscores the effectiveness of GAVI-supported immunization initiatives.

### 4.1 Socio-Demographic Characteristics (Table 1)

The socio-demographic characteristics of caregivers and children presented in Table 1 reveal critical insights into the population dynamics that influence immunization coverage and compliance in Bayelsa State. The data indicate a predominantly male-headed household structure, with 72.6% of caregivers being male. This aligns with traditional patriarchal norms prevalent in many Nigerian societies, where decision-making regarding health and welfare often falls to male household heads (Agbede et al., 2024). Such dynamics suggest that male caregivers may play a significant role in determining vaccination practices and could potentially influence the level of engagement in health-seeking behaviours, including immunization. The age distribution of children showcases a high proportion of infants under one year old, accounting for 44.3% of the sample. This demographic is particularly vulnerable to vaccine-preventable diseases (VPDs), highlighting the importance of timely immunization during this critical developmental stage (Wiysonge et al., 2012). The emphasis on vaccinating infants aligns with global health recommendations that prioritize early immunization as a means to mitigate the risks of morbidity and mortality associated with VPDs (WHO, 2019). The educational background of household heads is noteworthy, with a significant proportion having attained secondary education (54.8%) and 30.0% holding tertiary qualifications. Studies have shown that higher levels of parental education are positively correlated with improved health-seeking behaviours, including adherence to vaccination schedules (Dube et al., 2016). Educated caregivers are often better equipped to understand the importance of vaccines and are less susceptible to misinformation, which can significantly enhance vaccination rates (Agbede et al., 2024). This finding underscores the need for targeted educational initiatives aimed at lower-educated groups to bridge knowledge gaps and improve immunization uptake. The religious affiliation of the population is predominantly Christian (90.2%), which reflects the socio-cultural context of Bayelsa State. The influence of religion on health behaviours, including vaccination, has been documented in various studies, indicating that religious beliefs can either support or hinder vaccine acceptance (Agbede et al., 2024). Engaging local religious leaders in vaccination campaigns could foster a supportive environment for immunization and address potential vaccine hesitancy stemming from religious concerns. Additionally, the data reveal that 57.6% of household heads are self-employed, which may reflect economic conditions that influence access to healthcare services. The reliance on self-employment can pose challenges for families, as irregular incomes may affect their ability to prioritize health expenditures, including vaccinations (Bangura et al., 2020). This economic context reinforces the necessity for health policies that consider the financial barriers faced by families in accessing immunization services. The majority of the population resides in semi-urban areas (80.7%), which presents unique challenges regarding healthcare access. Geographic barriers have been linked to lower immunization coverage, particularly in semi-urban settings where health facilities may be less accessible compared to urban centres (Wiysonge et al., 2012). This finding emphasizes the importance of developing community-specific strategies that address the logistical challenges faced by families in accessing vaccination services. In summary, the socio-demographic characteristics outlined in Table 1 highlight the complex interplay of cultural, educational, economic, and geographic factors that influence immunization coverage in Bayelsa State. These insights are crucial for informing targeted interventions and educational initiatives aimed at improving vaccination rates among children, ultimately contributing to better public health outcomes in the region. Addressing the identified challenges through community engagement and tailored strategies will be essential to achieving universal immunization coverage and protecting vulnerable populations from vaccine-preventable diseases.

#### Vaccination Uptake (Figure 1)

The findings from this assessment of childhood vaccination uptake in Bayelsa State post-GAVI support demonstrate a substantial increase in the number of vaccinated children aged 0-59 months across various antigens. The data indicate that the overall uptake of vaccines rose significantly from 36,947 doses administered in the last quarter of 2021 to 86,301 doses during the same period in 2024. This remarkable growth can be attributed to the strategic interventions and funding provided by GAVI, which aimed to enhance immunization coverage in underserved settlements. The increase in vaccination for specific antigens reflects a positive trend in public health efforts. For instance, the BCG vaccine uptake was more than double from 6,447 to 16,407 doses, while measles vaccination rose from 6,947 to 11,361 doses. This increase is consistent with findings from other studies, which have shown that targeted funding and resource allocation can significantly enhance immunization rates in low-income countries (GAVI, 2018; WHO, 2025). The impressive uptake of oral polio vaccine (OPV-3) and pentavalent (Penta-3) vaccines, with increases from 7,962 to 20,324 and 7,964 to 20,317 doses respectively, further underscores the success of GAVI’s support in addressing vaccine-preventable diseases in the state. The substantial rise in yellow fever vaccinations, from 7,627 to 17,892, is particularly noteworthy given the disease’s potential for outbreaks in susceptible populations. This uptick can be seen as a proactive measure in disease prevention, aligning with global health initiatives aimed at eliminating yellow fever transmission (WHO, Elimination of Yellow fever Epidemics (EYE) Strategy 2017-2026). Such proactive measures are crucial in areas like Bayelsa State, where ecological factors can heighten the risk of transmission. However, while the data indicate an overall improvement in vaccination rates, it is essential to consider the underlying factors that contribute to these outcomes. Community engagement, health education, and improved healthcare infrastructure are pivotal in sustaining these gains (Abdullah et al., 2022). The role of local health workers in mobilizing communities and educating parents about the importance of vaccination cannot be over-emphasized. As noted by Xie et al., (2024), community communities, local trust, and accessibility significantly influence vaccine uptake, emphasizing the need for continued investment in community health initiatives. Moreover, the sustainability of these gains will depend largely on the ongoing commitment from both governmental, non-governmental organizations, and partners to maintain funding and support systems that ensure consistent vaccine availability and access. Long-term strategies should include routine monitoring and evaluation of vaccination programs to identify and address barriers to access, particularly in rural and remote areas. In the context of this conversation, the assessment of childhood vaccination uptake in Bayelsa State reveals significant progress following GAVI’s support. While the findings are encouraging, continued efforts are necessary to ensure that these gains are not only maintained but built upon, enhancing the resilience of the immunization program against future challenges. The lessons learned from this initiative can inform similar interventions in other regions facing comparable health challenges.

**Figure 1:**
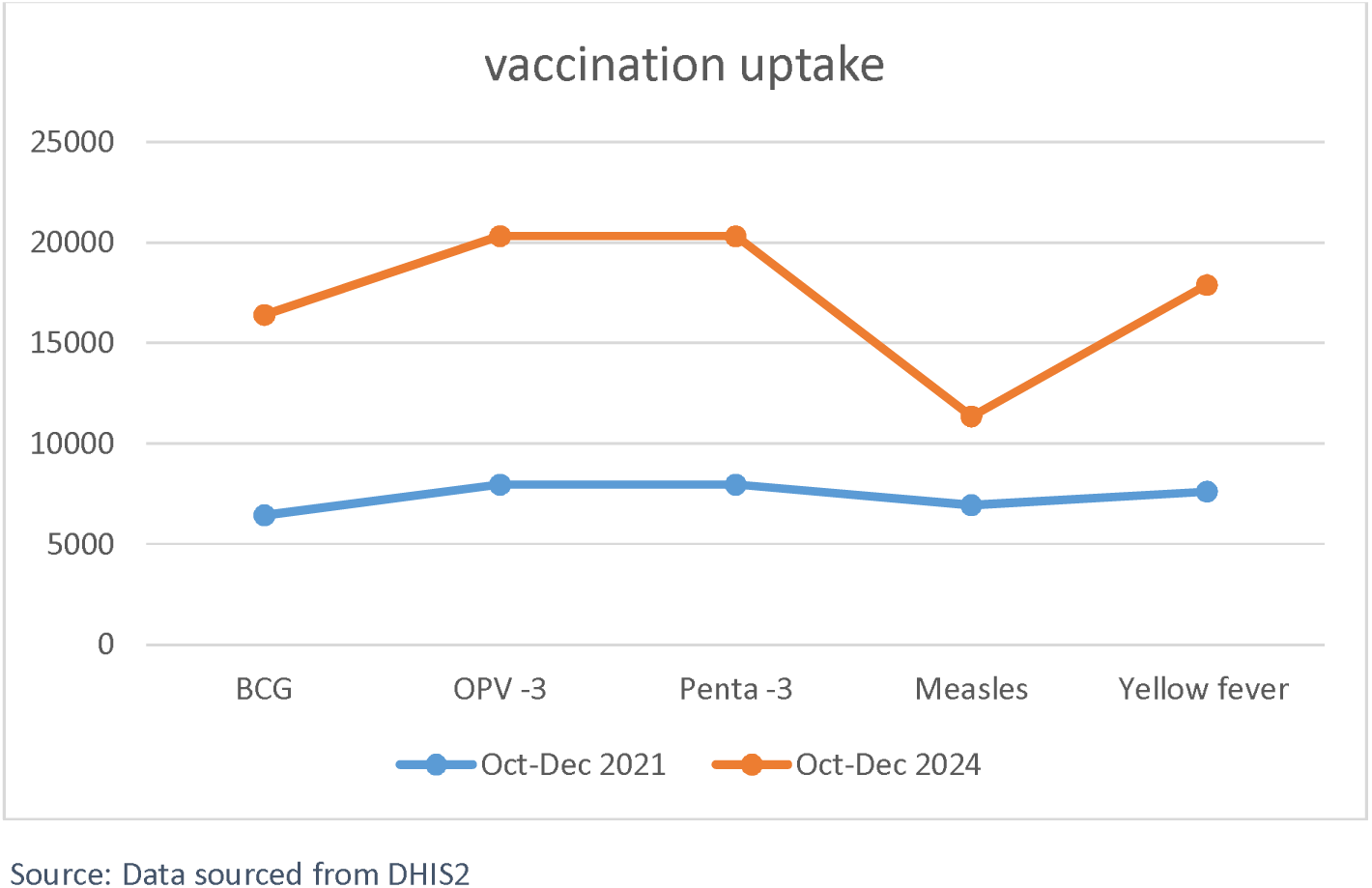
Vaccination Uptake

### 4.2 Immunization Coverage Rates by Antigen (Table 2)

The variability in immunization coverage rates across different antigens is noteworthy. While BCG and OPV achieved high coverage rates of 93.6% and 94.3%, respectively, the coverage for measles (56.0%) and yellow fever (54.5%) raises concerns. This pattern reflects trends observed in other studies, where initial vaccinations are often prioritized (Wiysonge et al, 2012). The lower uptake of the measles vaccine is particularly alarming, as it remains a highly contagious disease that can lead to severe complications. This discrepancy suggests the need for intensified health education and community engagement strategies, as proposed by Jibrin AM (2025), who identified caregiver misconceptions as a significant barrier to achieving comprehensive immunization. The literature indicates that caregivers may prioritize vaccines perceived as more urgent or necessary in the early stages of a child’s life, resulting in lower compliance for later vaccines, such as measles (MacDonald NE, 2015). Furthermore, the lower coverage rates for the measles and yellow fever vaccines could be attributed to misinformation and cultural beliefs, which have been documented as significant barriers to vaccination uptake (Agbede et al., 2024). In summary, while the immunization coverage for BCG and OPV is commendable, the lower rates for the measles and yellow fever vaccines signal a critical gap that needs immediate attention. This discrepancy underscores the need for targeted interventions, including enhanced community education and outreach efforts to address misconceptions and barriers to vaccination that can hinder compliance with the full immunization schedule. Such strategies are essential to mitigate the risk of vaccine-preventable diseases and achieve the WHO’s goal of 90% coverage across all antigens.

### 4.3 Overall Immunization Coverage (Table 3)

The overall immunization coverage of 95.5% is a remarkable improvement from the baseline coverage of 61.12% prior to the GAVI intervention. This substantial increase reflects the effectiveness of the GAVI-supported programs in enhancing vaccination uptake in Bayelsa State. The findings are consistent with other scholars, who found that strategic community engagement and targeted interventions significantly improve immunization rates in low- and middle-income countries. However, the observed variability in coverage among different vaccines underscores the necessity for sustained efforts to achieve uniformity across all vaccinations, particularly for those that are lagging, such as measles and yellow fever. This increase in immunization coverage aligns with findings from other studies that highlight the positive outcomes of targeted interventions and community engagement in vaccination programs. For instance, research by Wiysonge et al. (2012) emphasizes that strategic support and capacity-building efforts, such as those provided by GAVI, have been instrumental in elevating immunization rates in low- and middle-income countries, including Nigeria. Furthermore, the current findings resonate with the systematic review conducted by MacDonal, (2015), which identified that effective immunization programs result in improved vaccine coverage when they incorporate community engagement and address socio-cultural barriers. The study underscores the importance of understanding local contexts and facilitating open dialogue about vaccines, which can lead to increased acceptance and participation in immunization campaigns. The literature also suggests that while overall coverage is impressive, a closer look at specific vaccines reveals variability. For example, while coverage for BCG (93.6%) and OPV (94.3%) is robust, the rates for measles (54.5%) and yellow fever (54.5%) are notably lower. This discrepancy highlights a critical area for further intervention, echoing the concerns raised by Bangura et al. (2020) about the need for sustained efforts to ensure all vaccines are equally accessible and accepted within communities to prevent outbreaks of vaccine-preventable diseases. In summary, the overall immunization coverage of 95.5% in Bayelsa State is a commendable achievement indicative of the successful implementation of GAVI-supported programs. However, ongoing efforts must focus on addressing the lower coverage rates of specific vaccines and understanding the barriers that still exist among certain segments of the population, as highlighted by various scholars in the literature. This multifaceted approach is essential for achieving comprehensive immunization and ensuring the continued health and well-being of children in the state.

### 4.4 Caregivers’ Perception of Vaccine Safety and Necessity (Table 4)

The findings presented in Table 4 highlight significant perceptions among caregivers regarding vaccine safety and necessity, even among those who have accepted vaccination for their children. Notably, 19.1% of caregivers expressed the belief that vaccines are not safe, and a substantial 40.4% reported concerns about potential side effects. These apprehensions reflect a pervasive atmosphere of vaccine hesitancy, where acceptance does not equate to confidence. A considerable portion, 17.0%, deemed vaccines unnecessary, while 23.4% cited religious or philosophical beliefs against vaccination. These statistics indicate a complex landscape of caregiver attitudes that can significantly influence commitment to future vaccination. While a high percentage of caregivers have engaged in the immunization process, the existence of doubts and fears surrounding vaccine safety poses a risk to ongoing compliance. Research has shown that perceptions of vaccine safety are critical determinants of vaccination behaviours. For instance, Dube et al. (2016) emphasize that concerns about vaccine safety can lead to delayed or missed vaccinations, which ultimately jeopardizes the effectiveness of immunization programs. Similarly, Jibrin AM (2025) finds that misinformation and negative perceptions about vaccines can deter caregivers from maintaining adherence to immunization schedules, even among those who have initially accepted vaccines. The presence of misconceptions regarding vaccine safety highlights the need for targeted educational initiatives to address these concerns. Engaging community leaders and healthcare providers to deliver accurate and reassuring information about vaccines can help mitigate fears and enhance caregiver confidence. The findings underscore the importance of not only promoting vaccine acceptance but also fostering ongoing trust and understanding among caregivers. This is particularly crucial in the context of Bayelsa State, where cultural beliefs and misinformation can significantly shape health behaviours (Agbede et al., 2024). In summary, while the data reflects a commendable level of vaccination acceptance among caregivers in Bayelsa State, the underlying perceptions of safety and necessity warrant immediate attention. Addressing these perceptions through comprehensive education and community engagement strategies is essential to ensuring sustained commitment to vaccination. By focusing on building trust and dispelling myths, health programs can enhance the effectiveness of immunization efforts, ultimately safeguarding children’s health against vaccine-preventable diseases.

### 4.5 Reasons for Non-Compliance with Vaccination Schedules (Table 5)

The primary reasons for non-compliance, particularly “forgetting the next visit” (34.6%) and “concerns about vaccine side effects” (21.2%), point to gaps in caregiver awareness and engagement with immunization schedules. This finding resonates with Dube’ et al. (2016), who identified forgetfulness as a significant barrier to vaccination adherence. The implications of cultural beliefs and misinformation are also echoed in the literature. Addressing these concerns through reminder systems and community education could bridge the gap and enhance vaccination compliance (Bangura et al, (2020). Furthermore, “I did not receive adequate information about vaccination” was cited by 7.7% of respondents, reinforcing the need for targeted educational outreach. This finding aligns with the broader literature, including studies by Dube’ et al. (2016), which underscore the importance of health education in improving vaccine acceptance. Inadequate dissemination of information can perpetuate misconceptions, leading to increased vaccine hesitancy and non-compliance. Lastly, the reported reasons of “I did not have access to healthcare services” (3.8%) and “religious beliefs” (5.8%) further illustrate systemic barriers to vaccination. These findings resonate with Bangura et al. (2020), who noted that geographic and logistical challenges remain significant hurdles to immunization, particularly in underserved areas. The intersection of geographic access and cultural beliefs presents a complex challenge that requires multi-faceted interventions. In summary, the reasons for non-compliance with vaccination schedules among caregivers in Bayelsa State reflect a combination of forgetfulness, concerns about vaccine safety, cultural influences, lack of information, and access issues. These factors underscore the necessity for comprehensive educational initiatives and community engagement strategies that address both informational needs and cultural sensitivities, thereby promoting higher vaccination compliance and ultimately enhancing the health of children in the state.

### 4.6 Accessibility and Distance to Health Facilities (Table 6)

The relationship between the distance to health facilities and vaccination accessibility highlighted in Table 6 underscores a critical barrier to immunization efforts in Bayelsa State. The significant chi-square value (X²=10.84, p-value=0.001) indicates a strong association between proximity to healthcare services and vaccination compliance among children aged 0-59 months. Specifically, households located within 1 km of health facilities demonstrated higher vaccination participation rates compared to those living further away. This finding aligns with existing literature that emphasizes the detrimental impact of geographic barriers on healthcare access and service utilization. Distance to health facilities has long been recognized as a significant determinant of health service utilization, including immunization coverage (Wiysonge et al., 2012). Families residing in rural or remote areas often face logistical challenges that deter them from seeking timely vaccinations, such as transportation costs, time constraints, and physical barriers (Bangura et al., 2020). These challenges can exacerbate disparities in immunization rates, particularly in state where healthcare infrastructure is already limited. For instance, a systematic review by Dube’ et al., (2016) found that geographic barriers significantly contribute to lower immunization rates, particularly among disadvantaged populations. Similarly, a study conducted by (Sato R, 2020), in Nigeria reported that families living more than 5 km from health facilities were less likely to comply with vaccination schedules, highlighting the need for targeted interventions to improve access in these areas. In Bayelsa State, where considerable geographical diversity exists, it is imperative to devise strategies that enhance accessibility to immunization services. Implementing mobile vaccination units, community-based outreach programs, and increasing the number of healthcare facilities in underserved areas could significantly improve vaccination rates. Furthermore, engaging community health workers to provide education and support could alleviate some barriers related to awareness and logistical challenges (Utrilla et al., 2025). In summary, the findings from Table 6 not only confirm the critical role of geographic proximity in vaccination compliance but also emphasize the need for comprehensive strategies addressing accessibility issues. By improving physical access to healthcare services and leveraging community resources, immunization programs can enhance coverage and protect children from vaccine-preventable diseases.

### 4.7 Educational Level of Household Heads and Card Retention (Table 7)

The findings from Table 7 reveal a significant association between the educational level of household heads and the retention of immunization cards among children aged 0-59 months in Bayelsa State. The Chi-square test results indicate a strong correlation, with a Chi-square value of 17.92 and a p-value of less than 0.00002, suggesting that higher educational attainment is linked to better card retention rates. Specifically, caregivers with tertiary education exhibited the highest retention rates of immunization cards, whereas those with no formal education demonstrated the lowest retention rates. These findings align with existing literature that consistently highlights the positive correlation between caregiver education and health-related behaviours, including vaccination compliance and record-keeping (Dube’ et al., 2016; Agbede et al., 2024). Educated caregivers are generally more informed about the importance of immunization and the role of vaccination records in tracking their children’s health, which enhances their ability to adhere to vaccination schedules. This phenomenon has been observed in various studies, demonstrating that educational interventions can significantly improve health outcomes. The results underscore the critical need for targeted educational initiatives aimed at lower-educated caregivers. As indicated by Dube et al. (2016),

lower educational levels are often associated with higher rates of vaccine hesitancy and non-compliance due to limited access to accurate information regarding vaccinations. This is particularly relevant in the context of Bayelsa State, where misconceptions and cultural beliefs about vaccines can significantly impede vaccination efforts. Moreover, the implications of these findings extend beyond mere card retention. They suggest that enhancing educational outreach could lead to improved immunization compliance, ultimately benefiting children’s health outcomes. By equipping caregivers with knowledge about the importance of vaccination and the role of immunization cards, public health strategies can be more effectively tailored to meet the needs of communities in Bayelsa State. In summary, the significant association highlighted in Table 7 reinforces the importance of educational interventions in strengthening immunization programs. These insights call for the integration of educational strategies within immunization initiatives, which could ultimately contribute to higher immunization rates and better health outcomes for children in Bayelsa State and similar contexts.

### 4.8 Influence of Vaccination Information Sources (Table 8)

The findings presented in Table 8 highlight the significant association between the sources of vaccination information and participation in immunization programs among caregivers of children aged 0-59 months in Bayelsa State. The chi-square analysis indicates that different sources of information have varying degrees of influence on vaccination compliance, with a p-value of 0.025 signifying a statistically significant relationship. The data reveals that caregivers who received information about vaccines from healthcare workers exhibited the highest level of vaccination participation. This aligns with existing literature emphasizing the critical role of healthcare providers as trusted sources of information in promoting vaccination uptake. According to Dube et al. (2016), healthcare professionals are pivotal in addressing vaccine hesitancy and misinformation, as they can provide accurate, evidence-based information that alleviates concerns regarding vaccine safety and efficacy. Furthermore, a systematic review by Agbede and Oladipo (2024) supports the notion that effective communication by healthcare workers significantly enhances caregivers’ confidence in vaccines, ultimately leading to improved immunization rates. Conversely, the data shows that information from social media and informal sources, such as family and friends, correlated with lower vaccination participation rates. The reliance on these channels can perpetuate misinformation and fears surrounding vaccines, as highlighted by the findings of MacDonald et al. (2015), which indicate that misinformation spread through social media can lead to increased vaccine hesitancy. In this context, caregivers may be more susceptible to negative narratives about vaccination, which can influence their decision-making regarding immunization for their children. Additionally, the results suggest that community announcements and religious organizations also play a role in shaping perceptions about vaccination. Engaging these sources could be beneficial in reinforcing positive messaging about vaccines within communities. As noted by Bangura et al. (2020), community leaders and religious figures often hold significant sway in influencing health behaviours, particularly in regions where cultural beliefs and practices have a profound impact on healthcare decisions. The findings emphasize the need for targeted health communication strategies that leverage trusted information sources, particularly healthcare workers, to improve vaccination participation. Enhancing the capacity of healthcare providers through training and resources can empower them to effectively communicate the importance of vaccines and address concerns raised by caregivers. Furthermore, public health campaigns should aim to counteract misinformation prevalent on social media by providing accurate, easily accessible information about vaccines. In conclusion, the influence of vaccination information sources on immunization participation underscores the importance of strategic health communication in addressing vaccine hesitancy. By fostering collaborations with healthcare providers and community leaders, public health initiatives can enhance the dissemination of accurate information and ultimately improve vaccination coverage among children in Bayelsa State.

### 4.9 Z-Test Results for Comparison of Current Immunization Coverage with Baseline Coverage (Table 9)

The findings from this study provide critical insights into the immunization coverage, card retention, and factors influencing vaccine compliance among children aged 0-59 months in Bayelsa State, Nigeria, following the GAVI-funded immunization initiatives. The overall immunization coverage of 95.5% represents a substantial improvement from the baseline coverage of 61.12%, indicating the positive impact of targeted interventions and community engagement strategies implemented by the GAVI Alliance. This significant increase aligns with previous studies that have highlighted the effectiveness of strategic support in enhancing vaccination rates in low- and middle-income countries (Wiysonge et al., (2012). Despite the commendable overall coverage, the variability in immunization rates among different vaccines raises important concerns. Specifically, BCG and OPV achieved high coverage rates of 93.6% and 94.3%, respectively, while the measles vaccine coverage was notably lower at 56.0%, and yellow fever at 54.5%. These discrepancies reflect a trend observed in other studies where initial vaccinations are prioritized over subsequent ones (Agbede et al., 2024). The lower uptake of the measles and yellow fever vaccines signals a critical gap that requires immediate and focused interventions. This is particularly alarming given that measles remains a highly contagious disease with severe complications (Dube et al., 2016). Therefore, it is essential to implement enhanced community education and outreach efforts to address misconceptions and barriers affecting vaccination uptake. Moreover, caregiver perceptions regarding vaccine safety and necessity emerged as a significant barrier to full immunization compliance. Approximately 19.1% of caregivers expressed concerns about vaccine safety, while 40.4% highlighted fears of potential side effects. Such fears resonate with findings from Jibrin AM (2025), which identified misinformation and cultural beliefs as substantial obstacles to vaccination uptake. The presence of 23.4% of caregivers citing religious or philosophical beliefs against vaccination further complicates efforts to achieve comprehensive immunization coverage, indicating the need for culturally sensitive interventions that engage community leaders and religious figures to advocate for vaccination (Agbede et al., 2024). The study also revealed that distance to health facilities significantly influences vaccination compliance, with households located within 1 km of health centres demonstrating higher participation rates. The chi-square analysis (X²=10.84, p-value=0.001) underscores the importance of geographic accessibility in facilitating timely immunization. These findings are consistent with existing literature that emphasizes the detrimental impact of geographic barriers on healthcare access (Bangura et al., 2020). Therefore, strategies such as mobile vaccination units and community-based outreach programs are critical to enhancing access for families living in remote areas. Furthermore, the educational level of household heads was significantly associated with immunization card retention, as indicated by the chi-square test results (X²=17.92, p-value<0.00002). Higher educational attainment correlated with better retention rates of immunization cards, aligning with previous research that suggests educated caregivers are more likely to understand and prioritize vaccination schedules (Dube et al., 2016). This finding reinforces the necessity for targeted educational initiatives aimed at lower-educated caregivers to improve health-seeking behaviours and ensure effective record-keeping. In summary, while the overall immunization coverage in Bayelsa State has seen remarkable advancements, the study identifies critical areas for ongoing attention. Targeted interventions are essential to address the lower coverage rates for specific vaccines, combat caregiver misconceptions about vaccine safety, and improve geographic access to immunization services. By fostering community engagement and strengthening the role of healthcare providers in delivering accurate information, stakeholders can develop more effective immunization strategies tailored to the unique needs of the communities they serve. The calculated z-value of approximately 33.98 is significantly greater than the critical value of ±1.96 for a 95% confidence interval. Since the calculated z-value exceeds the critical z-value, we reject the null hypothesis, indicating that there has been a statistically significant increase in immunization coverage in Bayelsa State following the GAVI intervention. This result suggests that the GAVI-funded immunization program has had a substantial positive impact on vaccination rates among children aged 0-59 months in Bayelsa State, achieving a notable improvement from the baseline coverage of 61.12% to the current coverage of 95.5%. The GAVI-supported immunization services in Bayelsa State have led to significant improvements in childhood vaccination coverage, with the overall immunization rate rising dramatically from a baseline of 61.12% to an impressive 95.5%. This substantial increase indicates the effectiveness of targeted interventions and community engagement strategies implemented by the GAVI Alliance. High coverage rates were observed for key vaccines such as Bacillus Calmette-Guérin (BCG) and Oral Polio Vaccine (OPV), with coverage rates reaching 93.6% and 94.3%, respectively. Despite these gains, challenges remain, particularly concerning the lower uptake of essential vaccines like measles and yellow fever, which stand at 56.0% and 54.5%. Additionally, the program has emphasized the importance of caregiver education, as higher educational attainment among household heads correlates with better retention of immunization cards and improved compliance with vaccination schedules. The program also highlighted the critical role of geographic accessibility, revealing that families living closer to health facilities exhibited higher vaccination rates. Overall, the GAVI-supported initiatives not only enhanced vaccine availability and coverage but also underscored the need for ongoing educational efforts to address misconceptions about vaccine safety and necessity, ultimately aiming to sustain high immunization rates and protect children from vaccine-preventable diseases.

### 5.0 Conclusion

This study provides a comprehensive assessment of childhood immunization coverage, card retention, and associated factors among children aged 0-59 months in Bayelsa State, following the GAVI-funded immunization initiatives. The findings reveal a significant improvement in overall immunization coverage, rising to 95.5% from a baseline of 61.12%, indicating the positive impact of targeted interventions and community engagement strategies implemented by the GAVI Alliance. This achievement underscores the effectiveness of strategic support in enhancing vaccination rates in a challenging health landscape. Despite the overall success, the study identifies critical areas for ongoing attention. Notably, the lower coverage rates for specific vaccines, such as measles (56.0%) and yellow fever (54.5%), highlight persistent gaps that require targeted interventions. Caregiver perceptions regarding vaccine safety and necessity emerged as substantial barriers to full immunization compliance, with many expressing concerns about potential side effects and the need for vaccines. Educational interventions are essential to addressing these misconceptions and enhance understanding of the importance of vaccinations. Moreover, the significant association between educational attainment of household heads and immunization card retention emphasizes the need for health systems to incorporate educational strategies. Improving caregivers’ knowledge about vaccination processes can lead to better record-keeping and compliance. Geographic accessibility remains a challenge, as evidenced by the correlation between distance to health facilities and vaccination participation. Addressing logistical barriers through mobile vaccination units and community-based outreach programs can further enhance access to immunization services, particularly in underserved areas. In conclusion, while the study demonstrates remarkable advancements in immunization coverage in Bayelsa State, continued efforts to address barriers related to caregiver knowledge, cultural beliefs, and geographic access are crucial. By fostering community engagement and strengthening the role of healthcare providers in delivering accurate information, stakeholders can develop more effective immunization strategies. This holistic approach will not only sustain high vaccination rates but also protect children from vaccine-preventable diseases, ultimately contributing to improving public health outcomes in the state. The insights gained from this study serve as a valuable resource for policymakers and healthcare practitioners to refine immunization programs, ensuring they are responsive to the unique needs of the communities they serve.

## Recommendations

The GAVI-supported immunization program has demonstrated remarkable success in enhancing vaccination coverage in Bayelsa State, achieving an impressive increase from a baseline of 61.12% to 95.5%. To ensure that these gains are sustained, it is critical to develop a comprehensive plan that addresses the challenges faced in maintaining high immunization rates. The following recommendations are proposed, drawing on insights from the GAVI experience and recognizing the need for innovative and sustainable strategies within the context of limited financial resources and complex geographical terrains.

### Strengthening Community Engagement and Education

*Leverage Community Leaders*: Engage local leaders, including religious and traditional authorities, to advocate for vaccination and address misconceptions. These figures can play a pivotal role in influencing community attitudes toward immunization and enhancing acceptance.

*Tailored Educational Campaigns*: Develop culturally sensitive educational materials that address specific concerns regarding vaccine safety and efficacy. Use local languages and relatable contexts to improve understanding and encourage dialogue within communities.

*Utilize Existing Networks*: Capitalize on the established networks of community health workers, midwives, and mobilizers to disseminate information about vaccination schedules, safety, and the importance of completing immunization series. Regular workshops and community meetings can serve as platforms for discussions and clarifications.

### Implementing Reminder and Follow-Up Systems

*Automated Reminder Systems*: Establish systems for sending timely reminders to caregivers about upcoming vaccination appointments via SMS or phone calls. This can help mitigate issues of forgetfulness, which was identified as a significant barrier to compliance.

*Home Visits for Follow-Up*: Train community health workers to conduct house-to-house follow-ups for missed vaccinations. This personal touch can provide caregivers with both information and support, reinforcing the importance of adhering to vaccination schedules.

### Enhancing Accessibility to Vaccination Services

*Mobile Vaccination Units*: Deploy mobile vaccination units to reach remote and underserved communities, ensuring that families have easier access to immunization services. These units can be equipped with vaccines, essential supplies, and trained personnel to provide vaccinations on-site.

*Improving Health Facility Infrastructure*: Advocate for the establishment and enhancement of healthcare facilities in underserved areas. Collaborate with government and NGOs to secure funding for infrastructure improvements that promote better access to vaccination services.

*Transport Solutions*: Develop partnerships with local transport providers to facilitate easier access to health facilities for families living in hard-to-reach areas. Consider creating a community transport system that aids in transporting caregivers and children to vaccination sites.

### Sustaining Financial Management and Resource Allocation

*Innovative Financial Strategies*: Implement new accounting software and financial management practices that were successful during the GAVI program to ensure transparency and efficient allocation of funds. Regular financial audits and reviews can help maintain accountability.

*Diversification of Funding Sources*: Actively seek partnerships with local and international organizations, private sector sponsorships, and government grants to create a diversified funding base. Emphasizing the success of the GAVI program can attract potential funders interested in supporting immunization efforts.

*Community-Based Financing Models*: Explore community-based financing models that encourage local investment in health services. This could include small contributions from families or community groups that directly support vaccination programs.

### Monitoring and Evaluation Framework

*Regular Assessment of Immunization Programs*: Establish a robust monitoring and evaluation framework to continuously assess immunization coverage, compliance rates, and the effectiveness of interventions. Use data-driven insights to adapt strategies as needed.

*Feedback Mechanisms*: Create channels for caregivers and community members to provide feedback on vaccination services. This information can be invaluable for adjusting programs to better meet community needs and addressing concerns.

*Research and Adaptation*: Conduct ongoing research to identify emerging barriers and facilitators related to immunization. Adapting strategies based on new findings will ensure that interventions remain relevant and effective in addressing community needs.

### Capacity Building for Healthcare Workers

*Ongoing Training Programs*: Provide regular training and professional development opportunities for healthcare workers to enhance their capacity in delivering immunization services, addressing community concerns, and effectively communicating with caregivers.

*Engagement in Decision-Making*: Involve healthcare workers in the planning and evaluation of immunization strategies. Their insights and experiences can guide the development of practical and culturally appropriate interventions.

Sustaining the gains made during the GAVI-supported immunization program in Bayelsa State requires a multifaceted approach that prioritizes community engagement, accessibility, financial sustainability, and continuous monitoring. By implementing these recommendations, stakeholders can create a resilient immunization system capable of addressing the ongoing challenges posed by geographical and socio-cultural factors. This strategic framework will not only protect children from vaccine-preventable diseases but also ensure that the achievements of the GAVI program are built upon and reinforced in the years to come.

## Data Availability

The data collected for this study on childhood vaccine immunization coverage in Bayelsa State were gathered using Google Forms, a widely used online survey tool. The structured questionnaire was designed to capture various aspects, including caregiver demographics, immunization status, card retention, and perceptions regarding vaccination. Upon completion of the data collection phase, the responses were automatically compiled into a Google Sheets format, which was subsequently exported into Microsoft Excel for further analysis. The dataset is available upon reasonable request. Interested parties can contact the corresponding author via email to obtain access to the anonymized dataset. The data will be shared in accordance with ethical considerations and the privacy of the participants, ensuring that no personally identifiable information is disclosed.

https://docs.google.com/spreadsheets/d/1GIrV6cMXBMIpyhR1kWfdMMtjF8g40fKgQkhwp2EapiY/edit?usp=sharing

## Acknowledgement

I would like to express my sincere gratitude to all those who contributed to the successful completion of this research study. First and foremost, I would like to acknowledge the invaluable support of my data enumerators, whose dedication and hard work played a crucial role in gathering the data necessary for this research. Their commitment to accuracy and attention to detail ensured that the information collected was reliable and comprehensive. I also extend my heartfelt thanks to the respondents who participated in this study. Their willingness to share their experiences and insights provided essential data that contributed significantly to our understanding of childhood immunization coverage in Bayelsa State. Without their cooperation, this research would not have been possible. Additionally, I would like to express my appreciation to the Ethics Committee of the Bayelsa State Primary Health Care Board for their guidance and approval of the study protocol. Their commitment to upholding ethical standards in research ensured the protection of participants’ rights and welfare throughout the study. Finally, I would like to thank all my colleagues, mentors, and friends who offered their support and encouragement during the research process. Your contributions have been invaluable, and I am grateful for your assistance and encouragement. Thank you all for your support and belief in this important work.

## Authors’ Contributions

In this research work, the authors’ contributions are distinct and collaborative, reflecting their expertise and roles within the study:

**Ebiakpor Bainkpo Agbedi**: As the lead author and corresponding author, Dr. Agbedi contributed significantly in the overall conceptualization and design of the study. He was responsible for the development of the research framework, including the formulation of research questions and objectives. Dr. Agbedi also played a crucial role in data collection, analysis, and interpretation of results. His expertise in public health and statistics informed the methodological approach, ensuring that the study adhered to rigorous scientific standards. Additionally, he contributed to writing the manuscript and synthesizing the findings into actionable recommendations.

**Mordecai Oweibia:** As a co-author, Dr. Oweibia’s contributions were vital in the public health aspects of the study. He assisted in designing the survey instruments and interpreting the data related to immunization coverage and compliance. His knowledge of public health practices enriched the discussion section, particularly in addressing barriers to vaccination and proposing community-based interventions. Dr. Oweibia also contributed to the literature review, ensuring that the research was contextualized within existing public health frameworks.

**Christopher Peres Ekiyor:** Dr. Ekiyor, as a co-author, brought additional insights into community health outreach and engagement strategies. His involvement included assisting in the data collection process and providing qualitative insights from key informants, which added depth to the quantitative data. He contributed to analysing the results and discussing the implications for policy and practice within the context of Bayelsa State.

Collectively, the authors collaborated on the drafting and revision of the manuscript, ensuring that the research findings were clearly articulated and aligned with the objectives of the study. Their combined expertise in public health, statistical analysis, and community engagement strengthened the overall quality of the research, making it a valuable resource for stakeholders aiming to improve immunization coverage in Bayelsa State.

